# New compartment model for COVID-19

**DOI:** 10.1101/2022.12.27.22283962

**Authors:** Takashi Odagaki

## Abstract

Population is separated into five compartments for COVID-19; susceptible individuals (S), pre-symptomatic patients (P), asymptomatic patients (A), quarantined patients (Q) and recovered and/or dead patients (R). The time evolution of each compartment is described by a set of ordinary differential equations. Numerical solution to the set of differential equations shows that quarantining pre-symptomatic and asymptomatic patients is effective in controlling the pandemic. It is also shown that the ratio of non-symptomatic patients to the daily confirmed new cases can be as large as 20 and that the fraction of untraceable cases in new cases can be as large as 80%, depending on the policies for social distancing and PCR test.

## Introduction

COVID-19 is still prevailing over the world except for Africa after three years since the first outbreak in China in 2019^1,2^. Numerous case studies have been reported in these three years. According to the reported data, COVID-19 exhibits the following unusual characteristics:

1. Pre-symptomatic patients are infectious^3^.
2. Significant part of patients is asymptomatic and infectious^4-10^.
3. Infectious patients can be detected by Polymerase Chain Reaction (PCR) test^11^.
4. Symptomatic patients are supposed to be quarantined in a hospital or at home^12^.
5. Significant fraction of patients is untraceable^13^.

Therefore, it is important to distinguish symptomatic patients and oligo-symptomatic patients.

In the standard SIR (Susceptible-Infected-Removed) model^14^, the pre-symptomatic patients are assumed not to be infectious, and the quarantined patients are included in the infected compartment and thus infectious though they do not contact with susceptible people. The SIR model cannot provide any information about the relation between the daily confirmed new cases and the number of infectious patients. In order to overcome this deficiency of the SIR model, the SIQR model has been introduced^15,16^ in which the quarantined patients are considered as a compartment. It is shown that the peak position of the number of quarantined patients appears later than the peak position of the infected patients and that the total number of infected individuals can be estimated from the number of the daily confirmed new cases^16^.

The SIQR model, however, cannot provide any information on the number of untraceable patients who are supposed to be infected from asymptomatic patients since asymptomatic patients are not treated as a compartment. On the basis of an analysis of infection process of COVID-19, it has been shown that the onset ratio can be deduced from the fraction of untraceable patients^17^.

In order to obtain relations among daily confirmed new cases, the onset ratio, the fraction of untraceable patients and the total number of infectious people, I propose a new compartment model consisting of susceptible individuals(S), pre-symptomatic patients (P), asymptomatic patients (A), quarantined patients (Q) and removed individuals (R) which include recovered and dead patients. To be more accurate, the P and A compartments should be considered as pre-quarantine patients and infected patients at large, respectively. This model is a modified version of the SIQR model where the I compartment in the latter is separated into P and A compartments in the former. I present the time evolution of the epidemic and argue that the fraction of untraceable patients and the number of infectious individuals can be related to the number of daily confirmed new cases.

First, the basic ordinary differential equations are introduced and various quantities are related to observables. Then, numerical solution to the set of ordinary differential equations is presented and the time dependence of the fraction of untraceable patients and the ratio of the number of infectious patients to the daily confirmed new cases are given. Results are discussed at the end.

### SPAQR model

Elementary infection processes among the population of each compartment are given as follows: (1) A susceptible individual becomes a pre-symptomatic infectious patient with rate *β* after it contacts with infectious pre-symptomatic or asymptomatic patients. (2) Symptomatic patients are always quarantined. (3) Non-symptomatic patients are quarantined with rate *q* by PCR test. (3) Quarantined patients and asymptomatic patients are removed (recovered or died) by rate *γ*′ and *γ*, respectively. (4) Pre-symptomatic patients become either symptomatic patients or asymptomatic patients at rate *δ* and the incidence rate is denoted by *a*. Figure 1 shows schematically the SPAQR model.

**Figure 1.**
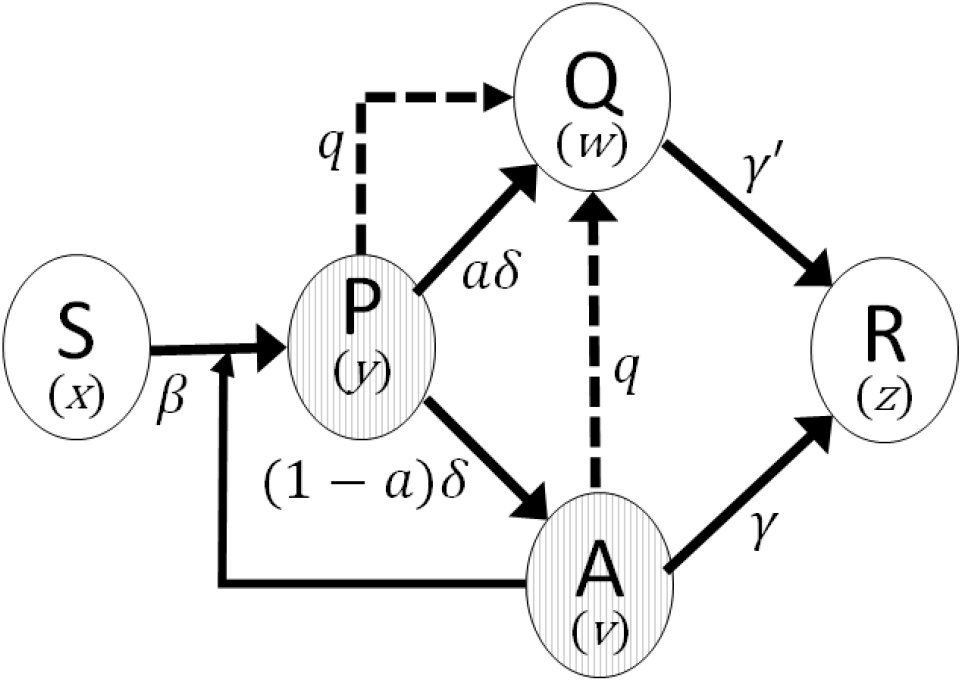
Schematic illustration of the SPAQR model for COVID-19.

The number of individuals in each compartment are denoted by *S, P, Q, A*, and *R*, and the total population is given by *N* = *S* + *P* + *A* + *Q* + *R*. Defining the fraction of individuals in each compartment by

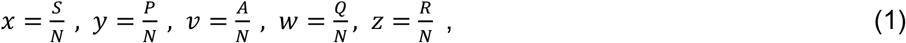

I assume that the time evolution of each compartment is governed by the following set of differential equations.

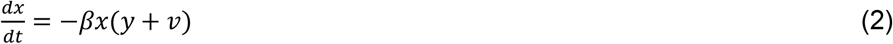

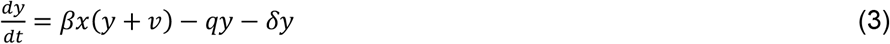

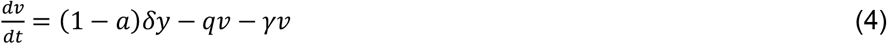

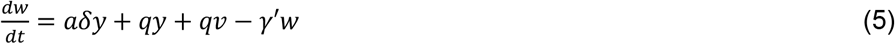

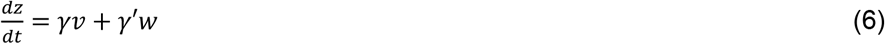

It is apparent that Eqs. (2)∼(5) satisfy the conservation of the population *x* + *y* + *v* + *w* + *z* = 1.

Besides the time dependence of each compartment, I am interested in observable quantities. First, the daily confirmed new cases Δ*Q* represent patients who are quarantined due to their symptom and who are tested positive though they are oligo-symptomatic, *i*.*e*. Δ*Q*/*N* = *aδy* + *qy* + *qv*. The number of infectious patients ℑ is given by ℑ/*N* = *y* + *v*. It is natural to assume that untraceable patients are infected from asymptomatic patients or pre-symptomatic patients who will not show any symptom later. Therefore, the fraction of untraceable patients *f* can be expressed as *f* = ((1 − *a*)*y* + *v*)/(*y* + *v*).

### Numerical simulation and results

The set of differential equations (2) ∼ (6) is stable and thus can be numerically solved by the Euler method. Setting *a* = 0.75, *δ* = 0.2, *γ* = 0.13, *γ*′ = 0.1^17^. I solved the set of equations (2) ∼ 6) for various *q* and *β*. The initial condition is set as *x* = 0.999, *y* = 0.001, *v* = *w* = *z* = 0.

Figure 2 show the time dependence of population in each compartment for (a) *q* = 0.0, *β* = 0.4, (b) *q* = 0.0, *β* = 0.3 and (c) *q* = 0.1, *β* = 0.4.

**Figure 2.**
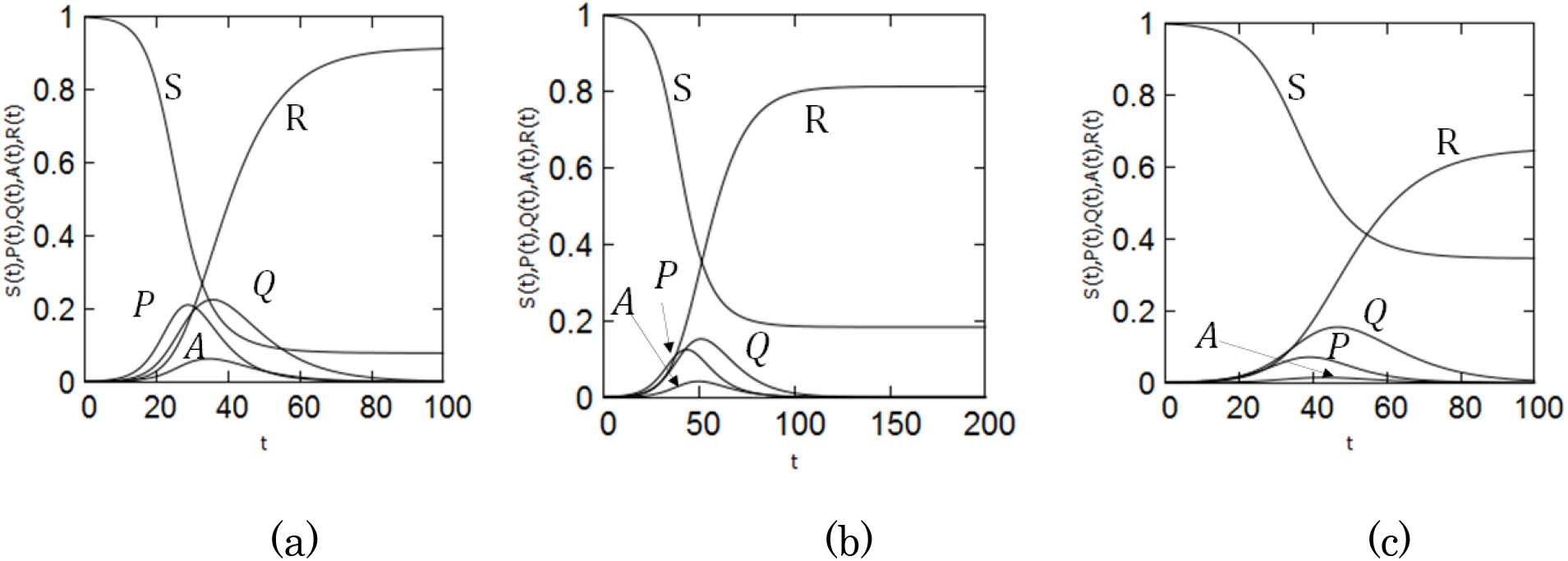
Time evolution of the population in each compartment of the SPAQR model for COVID-19. (a) *qq* = 0.0, *β* = 0.4, (b) *q* = 0.0, *β* = 0.3, (c) *q* = 0.1, *β* = 0.4.

The fraction of infectious patients is given by (P +A)/N = y + v, whose time dependence is shown in Fig. 3.

**Figure 3.**
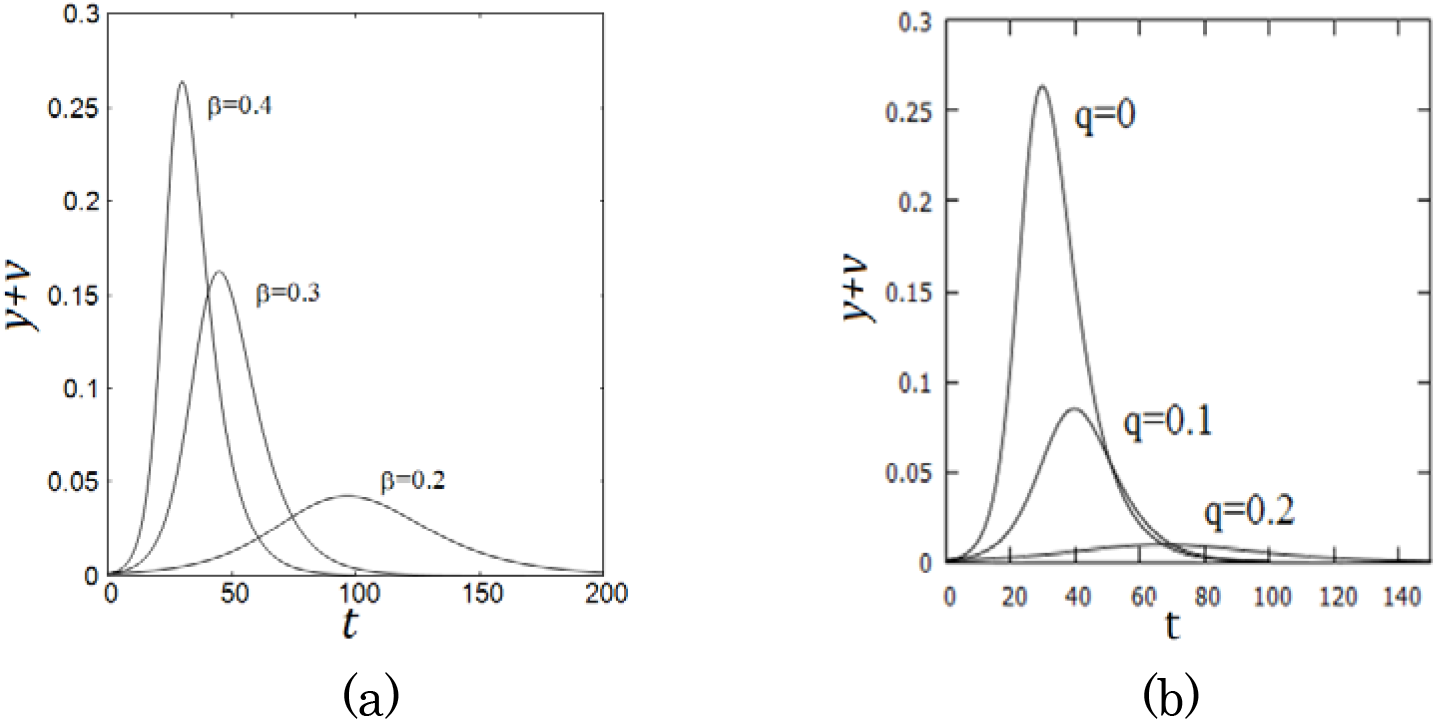
The time dependence of the number of infectious patients. (a) *β* = 0.4 0,3, 0.2 for *q* = 0.0 and (b) *q* = 0, 0.1, 0.2 for *β* = 0.4.

The daily confirmed new cases are given by Δ*Q*/*N* = *aδy* + *qy* + *qv* and thus the ratio of the number of infectious patients to the daily confirmed new cases is given by (*y* + *v*)/(*aδy* + *qy* + *qv*), which is shown in Fig. 4. When *a* = 0, the ratio is given 1/*q* as the SIQR model predicts^16^.

**Figure 4.**
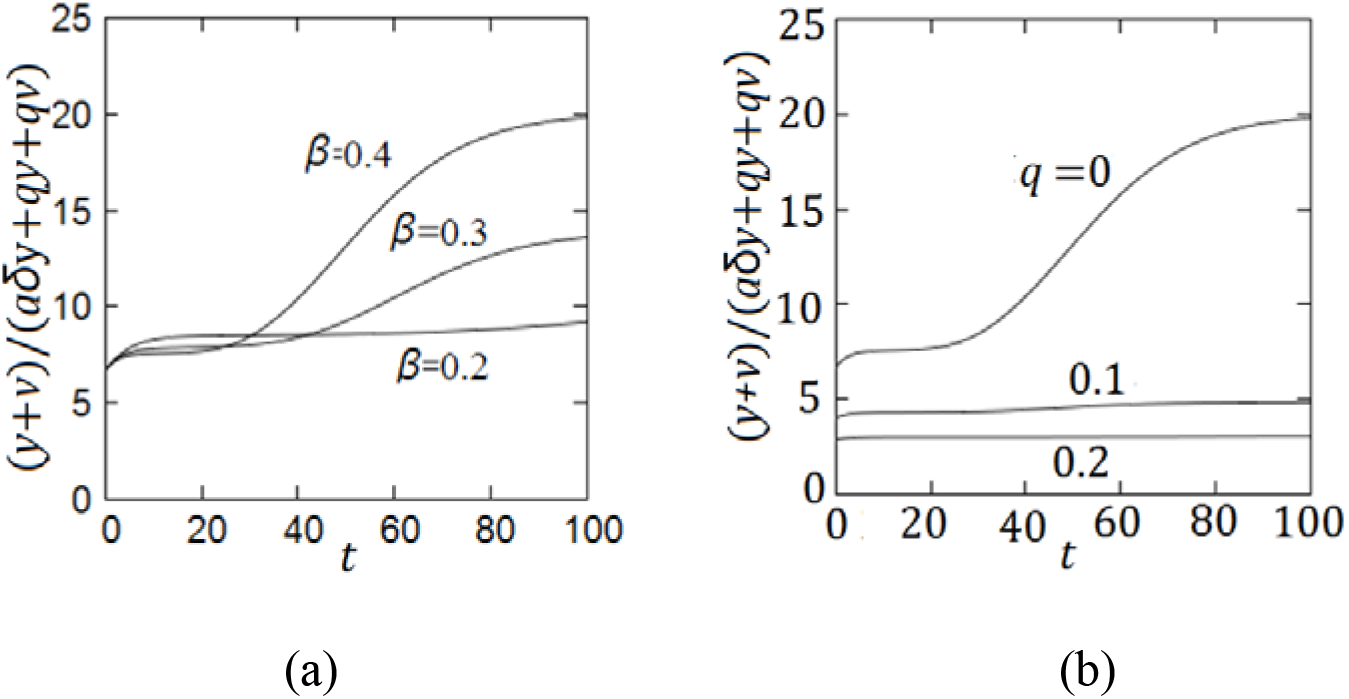
The ratio of infectious patients to the daily confirmed new cases is plotted as a function of time. (a) *β* = 0.4 0.3, 0.2 for *q* = 0.0 and (b) *q* = 0, 0.1, 0.2 for *β* = 0.4.

The fraction (1 − *a*) of pre-symptomatic patients will not show symptom after the incubation period, and individuals who are newly infected from these patients or asymptomatic patients are counted as untraceable patients. Therefore, the fraction of untraceable patients *f* in the daily confirmed new cases is given by *f* = ((1 − *a*)*y* + *v*)/(*y* + *v*) whose time dependence is shown in Fig. 5. In Tokyo, the fraction of untraceable patients was 60∼70% in 2022^13^, which is within the present estimation. Note that the onset rate *aa* can be estimated from the fraction of the untraceable patients^17^.

**Figure 5.**
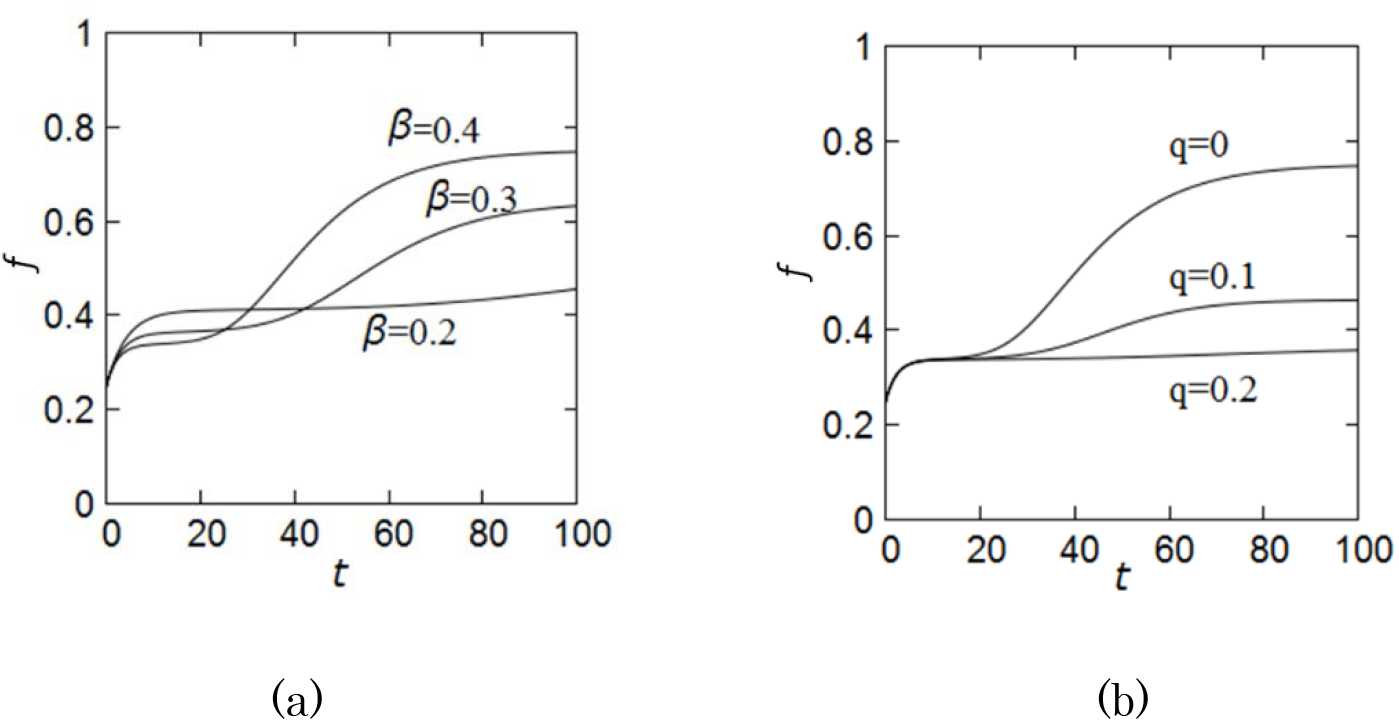
The fraction of untraceable patients in the daily confirmed new cases. (a) *β* = 0.4 0.3, 0.2 for *q* = 0.0 and (b) *q* = 0, 0.1, 0.2 for *β* = 0.4.

## Discussion

I have introduced the SPAQR model which will serve as an ultimate model for COVID-19.

Accumulated knowledges on COVID-19 imply that basic strategies controlling COVID-19 are; (1) quarantining pre-symptomatic and asymptomatic patients as well as symptomatic patients. (2) introducing strict social distance measure to reduce the transmission of the virus. (3) developing an effective medicine which removes the virus and (4) vaccination by a vaccine effective in prohibiting transmission of the virus. At the time when this paper was completed in 2022, the available vaccines are said not to be very effective in prohibiting the transmission of virus, and an effective medicine against the virus is yet to come, the present results strongly indicates that instead of so-called “with-CORONA policy” relying only on strategies (3) and (4) or so-called “zero-CORONA policy” relying strictly on strategy (2), a third policy using strategies (1) and (2) should be taken. Note that since infectious patients can be identified by PCR test, the lockdown measure need not to be enforced. except for the social distancing measure.

The fraction of infectious patients obeys

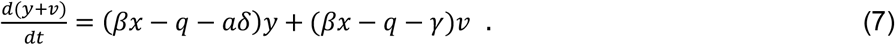

Consequently, the steady state where (*y* + *v*) = const can be realized regardless of the size of (*yy* + *vv*) since the condition is determined by the ratio *y*/*v*. Therefore, the policy for controlling COVID-19 must be formulated so that the fraction of infectious patients is as low as possible.

The parameters employed in the numerical simulation suggests that the fraction of untraceable patients is 60∼70% at the late stage of a wave^13^. This number is consistent with the observed value in Tokyo for the seventh wave, indicating that the parameter values are acceptable. As shown in Fig. 5, the number of infectious patients is about 20 times more than the daily confirmed new cases. Therefore, in order to control the epidemics, it is important to remove pre-symptomatic and asymptomatic patients by identifying them by PCR test.

## Data Availability

All data produced in the present work are contained in the manuscript

## Acknowledgements

This work was supported in part by JSPS KAKENHI Grant Number 18K03573.

## Data availability

The data supporting the findings of this study are available within the article.

